# Altered branched-chain α-keto acid metabolism is a feature of NAFLD in individuals with severe obesity

**DOI:** 10.1101/2022.02.07.22270618

**Authors:** Thomas Grenier-Larouche, Lydia Coulter Kwee, Yann Deleye, Paola Leon-Mimila, Jacquelyn M. Walejko, Robert W. McGarrah, Simon Marceau, Sylvain Trahan, Christine Racine, André C Carpentier, Aldons J. Lusis, Olga Ilkayeva, Marie-Claude Vohl, Adriana Huertas-Vazquez, André Tchernof, Svati H. Shah, Christopher B Newgard, Phillip J White

## Abstract

Hepatic *de novo* lipogenesis is influenced by the branched-chain α-keto acid dehydrogenase (BCKDH) kinase (BCKDK). We aimed to determine whether circulating levels of the immediate substrates of BCKDH, the branched-chain α-ketoacids (BCKAs) and hepatic *BCKDK* expression are associated with the presence and severity of non-alcoholic fatty liver disease (NAFLD).Eighty metabolites (3 BCKA, 14 amino acids, 43 acylcarnitines, 20 ceramides) were quantified in plasma from 288 bariatric surgery patients with severe obesity (BMI > 35 kg/m^2^) with scored liver biopsy samples. Metabolite principal component analysis (PCA) factors, BCKA, branched-chain amino acids (BCAA), and the BCKA:BCAA ratio were tested for associations with steatosis grade and presence of non-alcoholic steatohepatitis (NASH). Of all analytes tested, only the valine-derived BCKA, α-ketoisovalerate, and the BCKA:BCAA ratio were associated with both steatosis grade and NASH. Gene expression analysis in liver samples from two independent bariatric surgery cohorts showed that hepatic *BCKDK* mRNA expression correlates with steatosis, ballooning, and levels of the lipogenic transcription factor *SREBP1*. Experiments in AML12 hepatocytes showed that SREBP1 inhibition lowers *BCKDK* mRNA expression. These findings demonstrate that higher plasma levels of BCKA and hepatic expression of *BCKDK* are features of human NAFLD/NASH and identify SREBP1 as a transcriptional regulator of *BCKDK*.

## INTRODUCTION

Non-alcoholic fatty liver disease (NAFLD) is characterized by neutral lipid accumulation in the liver. In approximately one out of every five cases this is accompanied by pathologic inflammation and hepatocellular damage (ballooning), termed steatohepatitis (NASH) (1). This more pathogenic form of NAFLD progresses to fibrosis in approximately 35% of patients, significantly raising the risk for development of hepatocellular carcinoma (HCC), cirrhosis, and acute liver failure. Advanced NAFLD is also a significant risk factor for development of type 2 diabetes and cardiovascular diseases (CVD) (2, 3).

The obesity pandemic has driven a sharp increase in the incidence of NAFLD in recent years to an estimated level of 25% in the United States, and the incidence of NALFD-related liver failure is now comparable to hepatitis C as a primary cause of liver transplants (4). The propensity of an individual to develop NAFLD is dictated by a combination of genetics, lifestyle, diet, and insulin sensitivity (5, 6). Hepatic triglyceride pools are influenced by supply of adipose derived non-esterified fatty acids (NEFA) to the liver, hepatic *de novo* lipogenesis (DNL), NEFA export in very low-density lipoprotein (VLDL), and hepatic rates of beta oxidation and ketogenesis.

Importantly, metabolic flux studies show that high compared to low hepatic fat content in otherwise well-matched subjects with obesity is associated with three-fold higher rates of DNL but no difference in adipose efflux of NEFA or production of VLDL (7). Thus, hepatic DNL appears to be one important distinguishing feature of NAFLD status in the setting of obesity. Coupling of beta oxidation to the TCA cycle rather than ketogenesis may also be an underlying feature of persons with NAFLD (8), although this has yet to be studied in a population that is well matched for obesity but discordant for NAFLD.

We identified a new function for the branched chain α-keto acid dehydrogenase (BCKDH) kinase (BCKDK) and phosphatase (PPM1K) as post-translational regulators of ATP citrate lyase (ACLY), a key enzyme involved in hepatic DNL (9, 10). Whereas phosphorylation of BCKDH by BCKDK is inhibitory and promotes accumulation of BCKA and their cognate BCAA in plasma (11), phosphorylation of ACLY is activating and results in increased DNL via production of cytosolic acetyl CoA, which leads to the formation of the immediate DNL precursor and inhibitor of fatty acid oxidation, malonyl CoA (12, 13). Hepatic BCKDK levels are elevated in genetic models of obesity and following ingestion of diets high in fructose in rats, whereas PPM1K levels are low in these settings and increased during fasting (9, 10). Feeding of diets high in fructose induces expression of the lipogenic transcription factor Carbohydrate Responsive-Element Binding Protein-β (ChREBP-β), and ChREBP-β overexpression is sufficient to increase *BCKDK* and decrease *PPM1K* expression in rat liver (9). Also, adenovirus-mediated overexpression of recombinant BCKDK increases phosphorylation of ACLY and raises DNL by 2.5-fold in lean healthy rats (9). Conversely, inhibition of BCKDK with a small molecule, BT2, or adenovirus-mediated overexpression of recombinant PPM1K in liver of obese Zucker fatty rats lowers hepatic triglyceride content by >40%. These effects occur within seven days of BCKDK inhibition or PPM1K overexpression, in the absence of changes in food intake or body weight alongside robust lowering of circulating BCKA (9). We also found an association between hepatic *BCKDK* and *ChREBP*β expression in liver samples from humans with NASH (9), but the relationship of the metabolites directly regulated by this axis (i.e. BCKA) to NAFLD has not been examined.

Thus, we postulated that circulating levels of BCKAs correlate with NAFLD status in persons with obesity. We tested and validated this hypothesis in a well-characterized cohort of 288 bariatric surgery patients with severe obesity (BMI > 35 kg/m^2^) from the Quebec Heart and Lung Institute (QHLI) Obesity Biobank in whom liver biopsies were also taken at the time of bariatric surgery for histological grading of steatosis, inflammation, ballooning, and fibrosis. We also examined associations of hepatic *BCKDK* mRNA expression with hepatic steatosis, ballooning, and inflammation in two independent bariatric surgery cohorts. These studies highlighted a novel relationship between *BCKDK* and the lipogenic transcription factor Sterol Regulatory-Element Binding Protein-1 (SREBP1) that was subsequently explored *in vitro*.

## RESULTS

### Characteristics of the metabolite study population

To test our hypothesis that circulating levels of BCKA are associated with NAFLD status in obese persons we performed targeted metabolomic analysis in plasma samples from 288 bariatric surgery patients with severe obesity (BMI > 35 kg/m2) from the Quebec Heart and Lung Institute (QHLI) Obesity Biobank in whom liver biopsies were also taken at the time of bariatric surgery for histological grading of steatosis, inflammation, ballooning, and fibrosis. Demographic and clinical information for the study population is given in Table 1. A feature of the metabolite study population from the QHLI Obesity Biobank was that it allowed for stratification by steatosis grade or NASH status among subjects otherwise closely matched for age, gender, and BMI. With regards to clinical features associated with steatosis, we observed an increased prevalence of NASH and fibrosis in subjects with more severe steatosis. We also genotyped subjects for the presence of the Patatin-like phospholipase domain-containing protein 3 (*PNPLA3*) Ile148Met variant, previously shown to be commonly associated with elevated liver fat in Mexican-American subjects (14). At least one copy of Ile148Met variant, or G allele, was present in 48% of the patients studied here. Consistent with prior reports in Mexican-American subjects (14), the *PNPLA3* Ile147Met variant was more common in French-Canadian participants with higher steatosis grade; 27% of subjects with steatosis grade 0 had at least one copy of this variant vs. 78% of subjects with steatosis grade 3. Steatosis grade was also associated with impaired fasting glucose, HbA1c, insulin, and fasting plasma glucose levels. No association was found between steatosis grade and the proportion of individuals taking medications for blood pressure or lipids. Steatosis grade was also not associated with total cholesterol, high-density lipoprotein or low-density lipoprotein cholesterol concentrations, but was strongly associated with plasma triglycerides and circulating liver enzyme levels (ALT, AST, and GGT).

**Table 1.**
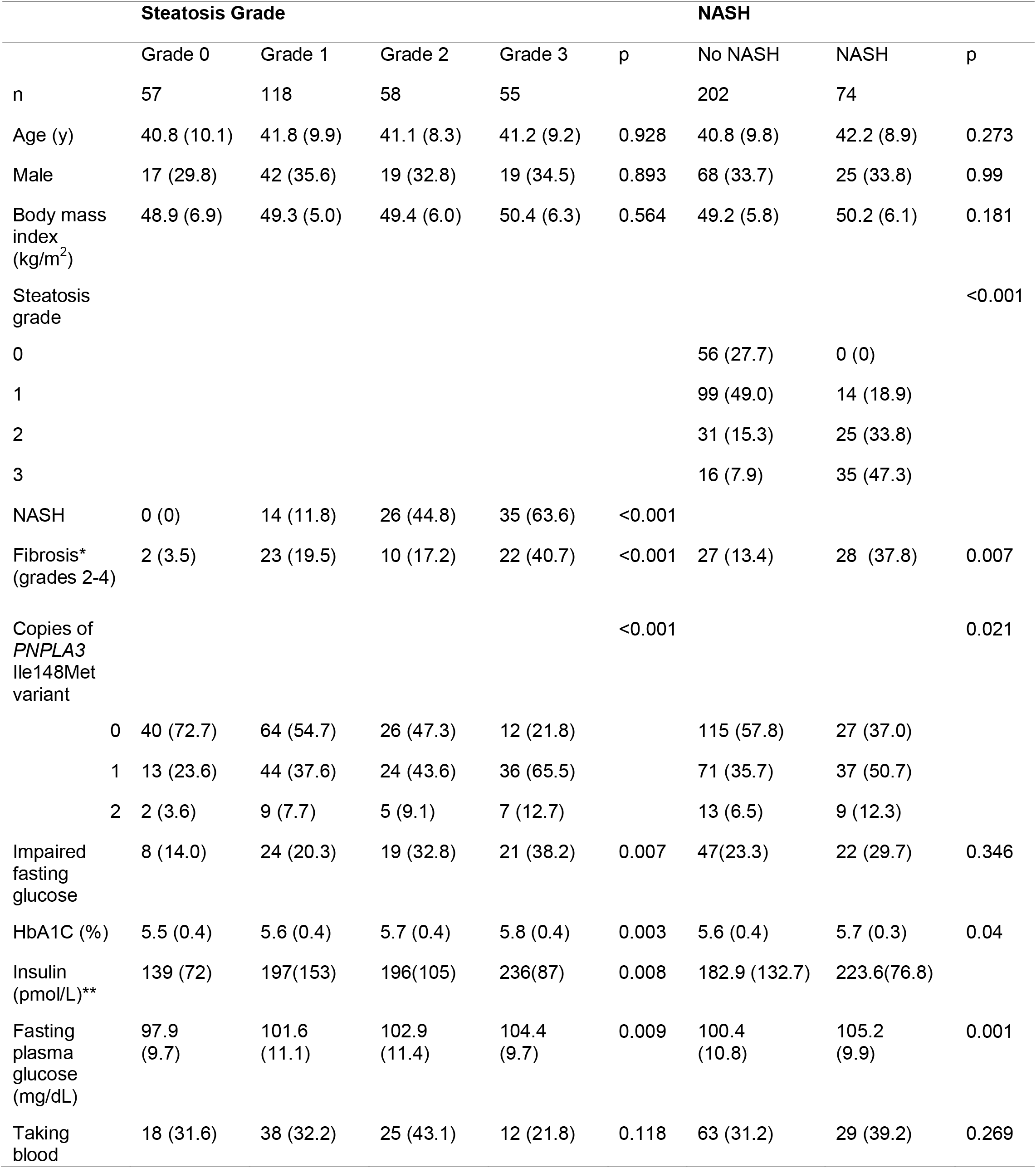
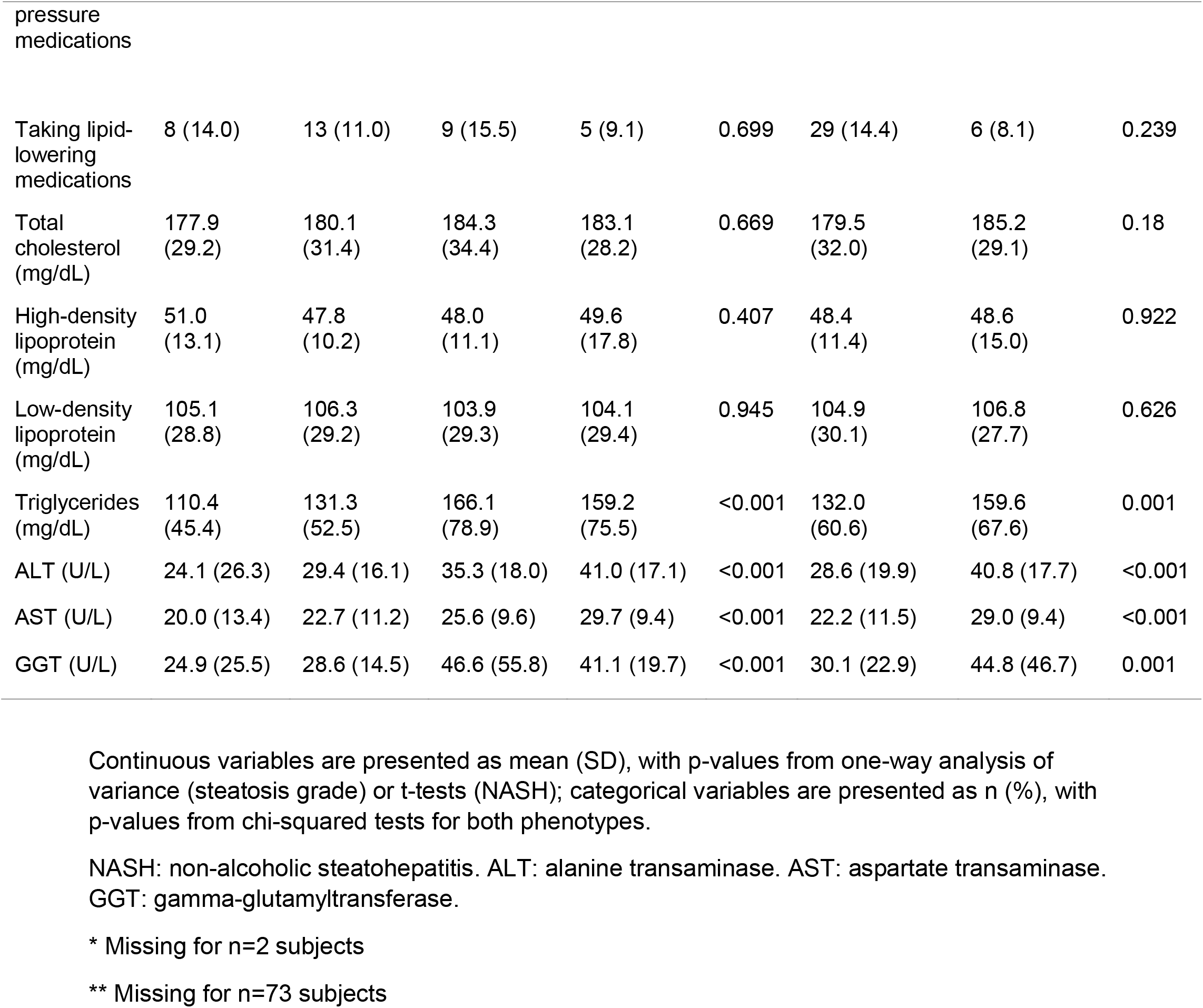
Baseline characteristics of the study population.

With regards to the clinical features of NASH, the presence of NASH was associated with more severe steatosis and individuals with NASH were more likely to have advanced fibrosis (grade 2-4) than those without NASH (38% vs. 13%). Associations between NASH and plasma lipids, liver enzymes, and medications mirrored those for steatosis grade. The *PNPLA3* Ile148Met variant, HbA1c, and fasting plasma glucose were also strongly associated with the presence of NASH.

### A BCKA-related signature of NAFLD status

In order to test the association between BCKA and NAFLD status in the broader context of other potentially relevant metabolic pathways we quantified 80 metabolites in the 288 plasma samples from the QHLI Obesity Biobank. The metabolites measured included the 3 BCKA, 14 amino acids, 43 acylcarnitines, and 20 ceramides. Principal components analysis (PCA) with varimax rotation was employed for dimensionality reduction of these 80 metabolites, resulting in 18 PCA factors with eigenvalue >1, explaining 73% of total variance (Table S1). We then tested associations of the three individual BCKA (KIV, KIC, KMV), molar sum of BCAA (Val, Ile/Leu), the BCKA:BCAA ratio (molar sum of KIV, KIC, and KMV divided by the molar sum of Val, Ile/Leu), and the 18 PCA factors with steatosis grade, NASH, and the presence of advanced fibrosis using univariate and multivariate regression.

In univariate analyses, only KIV (the BCKA derived from valine) and the standardized BCKA:BCAA ratio were strongly associated with both steatosis grade and NASH (Table 2). Specifically, higher levels of KIV and larger BCKA:BCAA ratios were associated with higher steatosis grade (OR [95% CI]=1.2 [1.1-1.3], p=1.1×10^−4^ and OR [95% CI]=1.5 [1.2-1.9], p=2.9×10^−4^, respectively) and increased risk of NASH (OR [95% CI]=1.2 [1.1-1.3], p=4.0×10^−4^ and OR [95% CI]=2.0 [1.5-2.7], p=3.0×10^−6^, respectively). This association does not appear to be driven by BCAA because none of the individual BCAAs or the PCA factor 5 (Table S1), which is primarily comprised of the three BCKA and their cognate BCAA, displayed an association with steatosis grade or NASH.

**Table 2.** Metabolites significantly associated with either steatosis grade or NASH.

| Phenotype | Metabolite | n | Univariate model |  | Multivariate model |  |
| --- | --- | --- | --- | --- | --- | --- |
|  |  |  | OR (95% CI) | p | OR (95% CI) | p |
| Steatosis grade | Factor 14 | 288 | 1.52 (1.23-1.89) | $1.3 \times 10^{-4}$ | 1.65 (1.31-2.11) | $3.6 \times 10^{-5}$ |
| | Factor 7 | 288 | 0.58 (0.47-0.73) | $2.5 \times 10^{-6}$ | 0.59 (0.46-0.75) | $2.6 \times 10^{-5}$ |
| | KIV | 288 | 1.16 (1.08-1.26) | $1.1 \times 10^{-4}$ | 1.16 (1.06-1.26) | $8.2 \times 10^{-4}$ |
| | BCKA/BCAA | 288 | 1.49 (1.2-1.85) | $2.9 \times 10^{-4}$ | 1.34 (1.06-1.7) | 0.015* |
| NASH | BCKA/BCAA | 276 | 2.01 (1.51-2.71) | $3.0 \times 10^{-6}$ | 2.15 (1.54-3.06) | $1.2 \times 10^{-5}$ |
| | KIV | 276 | 1.19 (1.08-1.31) | $4.0 \times 10^{-4}$ | 1.27 (1.13-1.44) | $1.1 \times 10^{-4}$ |
|  | Factor 10 | 276 | 0.61 (0.45-0.82) | 0.0014 | 0.57 (0.39-0.82)) | 0.0029* |

\*Result is not significant after Bonferroni adjustment for multiple tests
OR>1 indicates that increased levels of the metabolite (or metabolites heavily loaded on the PCA factor) are associated with higher steatosis grade or increased risk of NASH. Multivariate model is adjusted for HbA1c, ALT, AST, GGT, BMI, sex, age, *PNPLA3* genotype, and study phase.

Besides KIV and the BCKA:BCAA ratio, no other metabolite or metabolite factor was associated with both steatosis grade and NASH in univariate analysis. Moreover, of the 18 PCA factors listed in Table S1, only Factor 14 (alanine and proline) and Factor 7 (glycine, serine, and histidine), were associated with steatosis grade, whereas Factor 10 (C20:4, C22, C18:2-OH acylcarnitines) was the only PCA factor associated with NASH alone (Table S1). No metabolites or factors measured in the present study were found to be associated with the presence of advanced fibrosis.

The associations of KIV with steatosis grade and NASH, and the BCKA:BCAA ratio with NASH were maintained when a multivariate statistical model that considered BMI, sex, age, HbA1c, ALT, AST, GGT, *PNPLA3* Ile148Met genotype, and study batch was applied (Table 2).

However, the associations of BCKA:BCAA ratio with steatosis grade and Factor 10 with NASH were somewhat attenuated after adjustment for these clinical variables. To explore this further, we tested for two-way interactions between sex and *PNPLA3* Ile148Met genotype and these metabolites. The interaction of the BCKA:BCAA ratio with sex was marginally significant (p=0.05); sex-stratified analysis then revealed that BCKA:BCAA is associated with steatosis grade in females (OR [95% CI]=1.7 (1.2-2.3), p=0.002), but the association is completely absent in the male cohort (OR [95% CI]=1.0 (0.7-1.5), p=0.9) (Figure S1). Similarly, we observed an interaction between Factor 10 and the *PNPLA3* Ile148Met variant in the NASH analysis (p=0.003). In stratified analyses, there was no association of Factor 10 and NASH in carriers of the *PNPLA3* Ile148Met variant (G allele) (OR [95% CI]= 0.8 (0.5-1.4) p=0.5), while subjects without a copy of the variant showed an association (OR [95% CI]= 0.3 (0.1-0.6), p=7×10^−4^) (Figure S1).

### Association of hepatic *BCKDK* mRNA expression with features of NASH in persons with severe obesity

We next explored the relationship between *BCKDK* mRNA expression in hepatic tissue and the individual features of NASH, using liver biopsies from a smaller cohort of 60 bariatric surgery patients with severe obesity from the QHLI Obesity Biobank, as well as existing transcriptomics data from 107 bariatric surgery patients with severe obesity from the Mexican Obesity Surgery (MOBES) cohort (Table S2). Despite the potential environmental and ethnic differences present among the French Canadian and Mexican cohorts, the relationship between hepatic *BCKDK* mRNA expression and features of NASH was remarkably similar (Table 3). Results from both cohorts were consistent with the strong associations observed for KIV, and the BCKA:BCAA ratio with steatosis and NASH, showing significant positive associations between hepatic *BCKDK* gene expression and steatosis grade, ballooning, and NASH. There was no association between *BCKDK* expression and lobular inflammation in either study.

**Table 3.** Association of Hepatic *BCKDK* mRNA expression with NASH traits.

| Phenotype | QHLI cohort (n=60) |  |  | MOBES cohort (n=107) |  |  |
| --- | --- | --- | --- | --- | --- | --- |
|  | R | *p-val* | *p-val*<sup>a</sup> | R | *p-val* | *p-val*<sup>a</sup> |
| Steatosis | 0.29 | 0.03 | 0.03 | 0.34 | 0.001 | 0.001 |
| Ballooning | 0.42 | 0.004 | 0.003 | 0.44 | 6.1x10<sup>-6</sup> | 7.7x10<sup>-6</sup> |
| Lob Inflam | 0.16 | *n.s.* | *n.s.* | 0.15 | *n.s.* | *n.s.* |
| NASH | 0.37 | 0.01 | 0.02 | 0.36 | 3.0x10<sup>-4</sup> | 4.4x10<sup>-4</sup> |

p-val<sup>a</sup> adjusted for age, sex, and BMI

### The lipogenic transcription factor SREBP1 regulates *BCKDK* mRNA expression

Our prior work had identified the fructose sensing lipogenic transcription factor, ChREBP, as a mechanism linking BCKDK to the lipogenic process (9). In line with this prior observation, we observed a positive association between the *ChREBP*α and *BCKDK* transcript levels in the sequencing data from the MOBES cohort (R=0.33, P=6.4 × 10^−5^). In light of our observation of a strong association of circulating BCKA and hepatic *BCKDK* expression with NAFLD/NASH in persons with severe obesity we aimed to determine whether another major lipogenic transcription factor, sterol response element binding protein (SREBP1) might also regulate hepatic *BCKDK* gene expression. Examination of SREBP1 ChIP-seq data and histone acetylation patterns surrounding the *BCKDK* locus in HEPG2 cells in the ENCODE database revealed that SREBP1 peaks are present in both the *BCKDK* promoter and the enhancer region upstream of the *BCKDK* gene where ChREBP was previously found to bind (**Fig 1A**). Sequence alignment data show that in contrast to the enhancer region identified to be bound by ChREBP which is present in humans and rats but not mice, the promoter region bound by SREBP1 is conserved across all species (**Fig 1A**). In line with these ChIP-Seq data, examination of the relationship between *SREBP1* and *BCKDK* transcript levels in the RNA-seq data from the MOBES cohort revealed a robust positive correlation between *SREBP1* and *BCKDK* mRNA expression in human liver (R=0.45, P=1.1 × 10^−6^; **Fig 1B**). We therefore directly tested the regulation of *BCKDK* mRNA expression by SREBP1 in AML12 hepatocytes *in vitro* using two well validated chemical inhibitors of SREBP1, Betulin (15) and PF-429242 (16). 48 hours post treatment with either Betulin or PF-429242, AML12 hepatocytes displayed significantly lower expression of the SREBP1 target gene, fatty acid synthase (*FASN*), as well as *BCKDK* (**Fig 1C**). Together these data place SREBP1 alongside ChREBP as a lipogenic transcriptional regulator of *BCKDK* gene expression.

**Figure 1.**
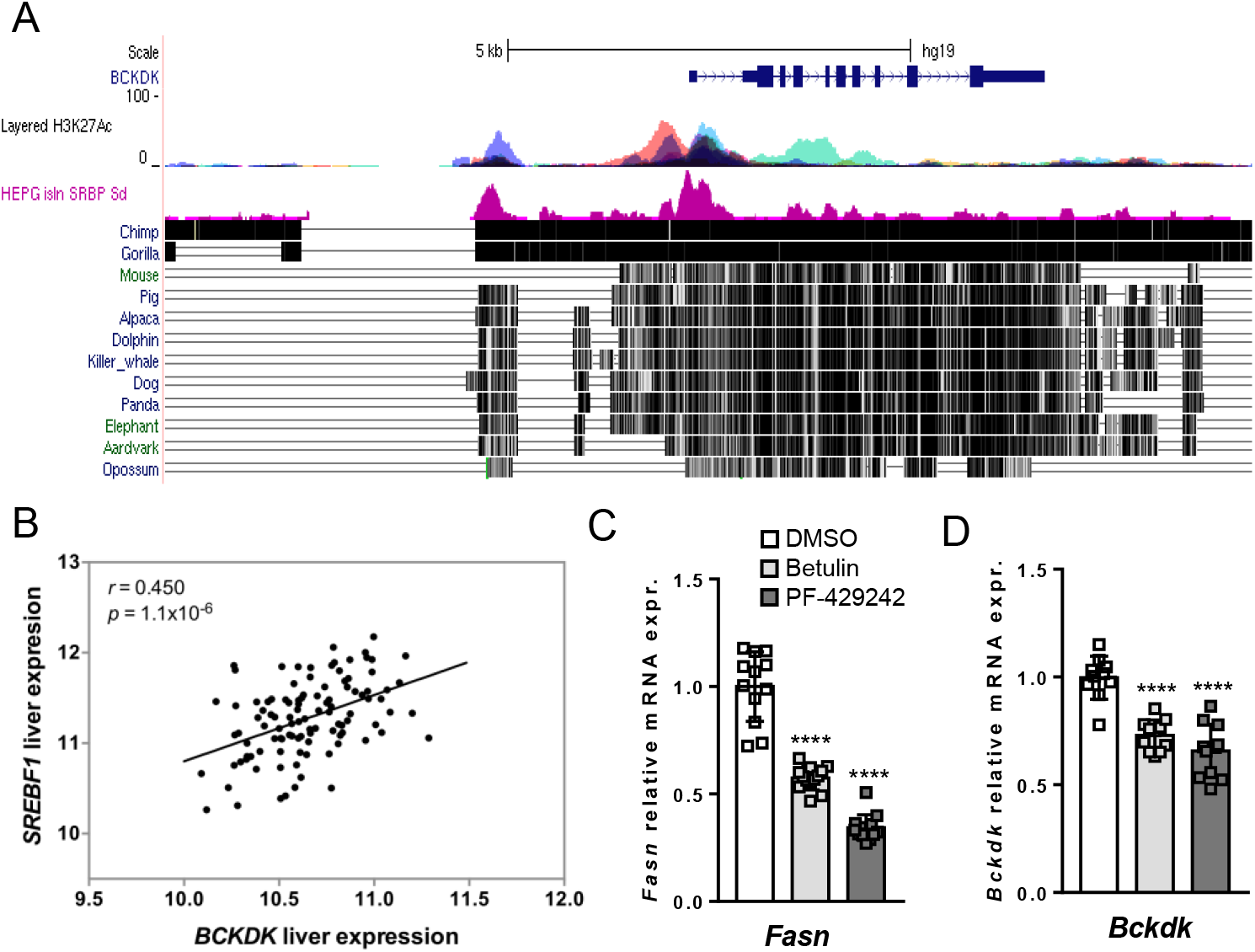
Regulation of *BCKDK* expression by SREBP1. Panel A shows H3K27Ac peaks (multicolored) and SREBP1 ChIP-Seq data (pink) in the promoter and upstream enhancer region of the *BCKDK* gene. Conservation of these genomic regions relative to the human genome is indicated by vertical black hatch marks to the right of each mammal. Panel B shows correlation between *BCKDK* and *SREBP1* gene expression in liver samples from the Mexican Obesity Surgery (MOBES) cohort. Panels C and D show effect of the SREBP1 inhibitors, Betulin and PF-429242, on *FASN* and *BCKDK* mRNA expression in AML12 cells. Data are expressed as the mean ± sem of three independent experiments. **** indicates P<0.001.

## DISCUSSION

Our metabolomic analysis of plasma from a well-characterized population of persons with severe obesity revealed that the BCKA, KIV, and the BCKA:BCAA ratio were the only metabolite features strongly associated with both steatosis grade and NASH even after adjustment for multiple covariables. Moreover, we demonstrated that the hepatic expression of *BCKDK* mRNA is strongly associated with steatosis grade, ballooning, and NASH in liver of two distinct bariatric surgery populations with severe obesity. These results are consistent with our finding in rats showing that pharmacologic or molecular manipulation of the hepatic BCKDK:PPM1K ratio to favor BCKDK simultaneously increases circulating BCKA levels, via inhibition of BCKDH, and hepatic DNL, via activation of ACLY (9). The current findings expand upon our studies in rodents to implicate this novel integrative metabolic regulatory node as a potential therapeutic target for human NAFLD. Our studies also introduce SREBP1 as a second lipogenic transcription factor, joining ChREBP-β, in regulation of *BCKDK* gene expression.

Finally, our data provide evidence for a potential clinical utility for plasma levels of KIV or the BCKA:BCAA ratio to identify individuals with NAFLD/NASH that might be best suited to therapies that target the lipogenic machinery.

BCAAs have repeatedly emerged as strong biomarkers of cardiometabolic disease traits such as obesity, insulin resistance, and future diabetes development (10, 17–21). More recently, a number of studies have demonstrated an association between BCAA and presence of NAFLD (22–26). Our work herein suggests that BCKA are a more sensitive indicator of NAFLD and NASH status than BCAA in persons with severe obesity. We propose that this is due to the fact that circulating BCAA can be impacted by a diverse set of metabolic adaptations in the obese milieu including insulin resistance, dietary changes, altered gut microbiota, and downregulation of components of BCAA catabolism in metabolic tissues other than liver such as adipose (10, 27–34). In contrast the liver does not metabolize BCAA due to very low levels of the branched-chain amino acid aminotransferase (BCAT) (35), whereas its high levels of BCKDH expression make it one of the most active sites of BCKA catabolism (36, 37). Thus, our data support the notion that measurement of plasma BCKAs or the ratio of BCKA:BCAA, which provides correction for systemic BCAA load, provides a sensitive index of the balance of BCKDK:PPM1K in the liver and NAFLD status.

Although the single BCKA, KIV, was strongly associated with both steatosis grade and NASH in both univariate and multivariate analyses, the association of the BCKA:BCAA ratio with steatosis grade was attenuated when the multivariate model was applied, and subsequently determined to be significant only in females. This sex interaction may be driven by differences in the partitioning of BCAA. Indeed, levels of BCAA and related metabolites are known to be higher in males than females, as shown in a cohort of adults who are overweight or obese (38), or in pediatric subjects with a comparable BMI (39). Moreover, BCAA levels were recently reported to be associated with NAFLD status in women but not in men (25). Moreover, sex-dependent differences in the relationship of BCAA with fasting glucose and lipids have been described in early adolescence (40). One possible mechanism underlying these observations is that the female sex hormone estrogen promotes BCAA uptake by inducing the expression of the cell polarity protein LLGL7L2, which binds to and activates the large neutral amino acid transporter SLC7A5 at the cell surface (41). However, additional work is warranted to better understand the differential regulation of BCAA utilization across the sexes and whether this contributes to any differences in risk for NAFLD and/or other cardiometabolic diseases.

Beyond KIV and the BCKA:BCAA ratio, we identified two additional amino acid-related factors associated with steatosis grade, as well as a long-chain acylcarnitine-related factor that was associated with the presence of NASH in carriers of the major *PNPLA3* allele. Factors 7 and 14, are made up of glycine related amino acids (glycine, serine, and histidine) and nitrogen handling metabolites (alanine and proline), respectively. Importantly, the negative association of glycine and serine with steatosis grade and BCAA levels observed here has also been reported in other cohorts (27, 37, 42–44). Factor 10, comprised of C20:4, C22, C18:2 carnitines, was found to associate with the presence of NASH, in univariate analysis. Given that the metabolite with strongest loading in this factor is arachidonyl (C20:4)-carnitine, it is tempting to speculate that the association between this factor and NASH is driven by arachidonic acid derived lipid mediators known to play a role in inflammation such as the leukotrienes and prostaglandins (45).

In conclusion, the current study provides proof of concept in humans that plasma levels of BCKA or the BCKA/BCAA ratio associate with NAFLD status in obese individuals. This finding also provides support for the idea that excessive hepatic expression of the BCKDH kinase, now understood to also play a role in regulation of the critical DNL enzyme ACLY (9), plays an important role in the development of NAFLD in human obesity. The major strengths of our study include the use of a well-characterized population matched for several confounders across grades of liver steatosis, the use of samples in which liver phenotypes were determined via gold standard histological grading, and validation of gene expression studies across obesity cohorts with different ethnic backgrounds. Future studies are warranted to verify if the associations described herein are also observed in other populations with different ethnic backgrounds and other forms of fatty liver disease.

## METHODS

### Study participants

The study population employed for plasma metabolomic analysis consisted of a cohort of patients of European ancestry with severe obesity (BMI > 35 kg/m^2^) from the eastern provinces of Canada who underwent bariatric surgery at the Institut universitaire de cardiologie et de pneumologie de Québec (Québec Heart and Lung Institute, QHLI, Québec City, QC, Canada). Of patients with available samples in the QHLI Obesity Biobank, 404 met the initial inclusion criteria for the study which were: HbA1c < 6% and fasting plasma glucose < 126 mg/dL, histologic NAFLD characterization, consent for genetic studies, and not on diabetes medications. NAFLD was present in 79% of these patients (steatosis grade 1 [5-33%]: 55%, grade 2 [34-66%]: 18%, grade 3 [>67%]: 6%). For this study 288 participants were used for the study were selected across ranges of steatosis grades (0-3) and steatosis with NASH (defined as the presence of steatosis alongside both lobular inflammation and ballooning) vs no NASH, matched on age, sex, BMI, and glucose tolerance. Evaluation of liver histology was performed by a pathologist according to Brunt *et al*. (46). Bio specimens were obtained from the Biobank of the Institut universitaire de cardiologie et de pneumologie de Québec according to institutionally-approved management modalities.

For liver gene expression analyses, a set of liver biopsy specimens from 60 individuals with severe obesity (BMI > 35 kg/m^2^) were also obtained from the QHLI Obesity Biobank. Similar to the samples used for metabolomic analysis patients were of European ancestry from the eastern provinces of Canada with severe obesity who underwent bariatric surgery at the Québec Heart and Lung Institute (QHLI, Québec City, QC, Canada) and were discordant for NAFLD and NASH as determined by a pathologist according to Brunt *et al*. (46). The results from this targeted gene expression analysis were validated using existing transcriptomic data from the Mexican Obesity Surgery (MOBES) cohort (47–49).

### Genotyping

Patatin-like phospholipase domain-containing protein 3 (*PNPLA3*) genotyping for the Ile148Met variant associated with hepatic steatosis (rs738409) was performed for samples from the QHLI Obesity Biobank on genomic DNA extracted from the blood buffy coat using the GenElute Blood Genomic DNA kit (Sigma, St. Louis, MO, USA). rs738409 was genotyped using validated primers and TaqMan probes (Applied Biosystems). PNPLA3 genotypes were determined using 7500 Fast Real-Time PCR System (Applied Biosystems).

### Metabolite profiling

BCKA, amino acid, acylcarnitine, and ceramide levels were measured in plasma by targeted metabolomics methods, as previously described (37, 50). Briefly, plasma concentrations of the alpha-keto acids of leucine (α-keto-isocaproate, KIC), isoleucine (α-keto-β-methylvalerate, KMV) and valine (α-keto-isovalerate, KIV) were measured by LC-MS and amino acid (n=15), acylcarnitine (n=45), and ceramide (n=21) profiling was performed by tandem mass spectrometry (MS/MS). All MS analyses employed stable-isotope-dilution with internal standards from Isotec, Cambridge Isotopes Laboratories, and CDN Isotopes.

### Gene expression analysis

For the human liver samples from the QHLI Obesity Biobank, PCR was used to measure gene expression. Specifically, total RNA was isolated using the total RNA purification kit (NORGEN BIOTEK). Total RNA for *in vitro* experiments was isolated using TRI-Reagent® (Sigma-Aldrich, #T9424). cDNA was generated using the high capacity reverse transcription kit (Applied Biosystems) and real-time qPCR was performed on a QuantStudio 6 Flex Real-Time PCR system (Applied Biosystems) using specific primers (Table S3) and the PowerUp SYBR Green Master Mix (Applied Biosystems), following the manufacturer’s instructions. Gene expression was normalized to RPLP0 using the ddCT method.

### RNA sequencing

The RNA isolation and sequencing for the MOBES cohort has been described previously (48, 49, 51). Briefly, sequencing was performed using an Illumina HiSeq2500 instrument. After data quality control, sequencing reads were mapped to the human reference genome using TopHat software v2.0.1 (52) and quantified using Cufflinks software (53).

### ChIP-seq analysis in ENCODE

The presence of SREBP1 ChIP peaks near the *BCKDK* transcription start site and previously identified ChREBP binding site upstream of the *BCKDK* gene was assessed using the HEPG2 SREBP1 Standard insulin ChIP-seq Signal from the ENCODE/SYDH data set, available in the ENCODE project with GSM935627 as the GEO sample accession number (54).

### AML12 experiments

Alpha mouse liver cells (cat. no. CRL2254; ATCC) were cultured in Ham’s F-12 (Gibco, ThermoFisher Scientific, #11320033) supplemented with 10% FBS (Gibco, ThermoFisher Scientific, #10437036), 1% glutamine (Gibco, ThermoFisher Scientific, #25030-081), 10 μg/mL insulin (Gibco, ThermoFisher Scientific, #12585014), 5 μg/mL transferrin (Sigma-Aldrich, #T1147), 5 ng/mL selenium (Sigma-Aldrich, #S9133), 40 ng/mL dexamethasone (Sigma-Aldrich, #D4902) and were maintained in a humidified incubator at 37°C under 5% CO2. Cells were plated in 24-well plates at a density of 80,000 cells/well. The next day, cells were treated with either 10 μM Betulin (Cayman chemical, #11041) (15), 10 μM PF-429242 (Cayman chemical, #15140) (16), or DMSO in complete growth media for 48h prior to harvesting of mRNA in TRI-reagent® for gene expression analysis.

### Statistics

For the primary study of metabolomics in the samples from the QHLI Obesity Biobank, 80 metabolites met quality control standards and were used in principal components analysis (PCA) followed by varimax rotation; 18 independent PCA factors with eigenvalue >1 were retained for subsequent analysis and explained 73% of the total variance. Each PCA factor is described by its primary metabolites (|loading|>0.4) in Table S1. In addition to PCA factors, we considered the three individual BCKA (KIV, KIC, KMV), BCAA (valine [Val], isoleucine/leucine [Ile/Leu]), and the BCKA to BCAA ratio (molar sum of KIV,KIC, KMV/ molar sum of Val, Ile/Leu, standardized to give a quantity with mean=0 and standard deviation=1). Proportional odds logistic regression was employed to test the association of the aforementioned variables with steatosis grade (Grade 0 (n=57), Grade 1 (n=118), Grade 2 (n=58), Grade 3 (n=55)). Logistic regression was used to test the metabolite/PCA factor associations with NASH (yes (n=74)/no (n=202)) and advanced fibrosis (Grade 0-1 (n=230) vs Grade 2-4 (n=57)). All associations were tested in both univariate models and multivariate models adjusted for HbA1c, liver enzyme levels (ALT, AST, GGT), BMI, sex, age, *PNPLA3* genotype, and technical batch. Associations were considered significant at p<0.0024, given a Bonferroni adjustment for multiple tests. Metabolites/factors that were significantly associated with a phenotype in univariate, but not multivariate, models were tested for interactions with sex or genotype, followed by stratified analyses as appropriate. Statistical analyses were carried out in R v4.1.2.

For gene expression analyses in both the QHLI cohort and the MOBES cohort, a partial Pearson’s correlation analysis corrected for age, sex, and BMI was used to assess the association between *BCKDK* mRNA expression and the individual features of NASH. A nominal P<0.05 was considered significant. Correlation between *BCKDK* and *SREBP1* expression within the MOBES transcriptomics data set was also assessed using a partial Pearson’s correlation.

The effects of the SREBP1 inhibitors Betulin and PF-429242 on *BCKDK* and *FASN* mRNA expression in AML12 cells were assessed using one-way ANOVA with a Dunnett’s post-hoc test, data represent three independent experiments and are expressed as mean ± the standard deviation. A nominal P<0.05 was considered significant.

### Study approval

All studies were conducted according to the principles outlined in the Declaration of Helsinki and approved by the Institutional Review Boards at Université Laval, UCLA, INMEGEN, and Duke University. All participants provided written, informed consent.

## Data Availability

All data produced in the present study are available upon reasonable request to the authors

## AUTHOR CONTRIBUTIONS

PJW, TGL, and CBN conceptualized studies. PJW, CBN, SHS, AHV, LCK, YD, LPM, RWM, and TGL interpreted data. PJW, TGL, CBN, SHS, AT, AHV, AJL, RWM, and ACC planned studies. LCK, PLM, JMW, and YD performed statistical analyses. SM, ST, and CR acquired samples. OI conducted metabolomics analyses, MCV performed PNPLA3 genotyping. PLM analyzed sequencing data. YD conducted AML12 studies. PJW wrote the manuscript. CBN, SHS, LCK, TGL, AT, MCV, and PLM edited the manuscript. All authors read and approved the manuscript in its final form. Order of co-first authors was determined by seniority on the project.

## ACKNOWLEDGEMENTS

The Authors acknowledge the invaluable collaboration of the surgery team, bariatric surgeons and biobank staff of the IUCPQ/QHLI and the participants of the studies. We thank Hugo Villamil and Samuel Canizales for sharing MOBES liver gene expression data. This work was supported by an American Diabetes Association Pathways to Stop Diabetes Initiator Award 1-16-INI-17 to PJW and Cardiovascular-Metabolic Fellowship 1-21-CMF-005 to JMW. NIH grants DK58398, DK121710, and DK124723 to CBN, NIH grant HL127009 to SHS, American Heart Association grant 17SFRN33670990 to SHS, a sponsored research agreement from Pfizer to CBN, and a Michael Smith Foreign Study Scholarship from the Canadian Institute of Health Research to TGL. MCV is Tier 1 Canada Research Chair in Genomics Applied to Nutrition and Metabolic Health.

## REFERENCES

1. Anstee QM, Reeves HL, Kotsiliti E, Govaere O, Heikenwalder M. From NASH to HCC: current concepts and future challenges. Nat. Rev. Gastroenterol. Hepatol. 2019;16(7):411–428.

2. Adams LA, Anstee QM, Tilg H, Targher G. Non-alcoholic fatty liver disease and its relationship with cardiovascular disease and other extrahepatic diseases. Gut 2017;66(6):1138–1153.

3. Salah HM et al. Relationship of Nonalcoholic Fatty Liver Disease and Heart Failure With Preserved Ejection Fraction. JACC. Basic to Transl. Sci. 2021;6(11):918–932.

4. Younossi Z et al. Global burden of NAFLD and NASH: trends, predictions, risk factors and prevention. Nat. Rev. Gastroenterol. Hepatol. 2018;15(1):11–20.

5. Friedman SL, Neuschwander-Tetri BA, Rinella M, Sanyal AJ. Mechanisms of NAFLD development and therapeutic strategies. Nat. Med. 2018;24(7):908–922.

6. White PJ, Abdelmalek MF. Insights Into Metabolic Mechanisms and Their Application in the Treatment of NASH. Clin. Liver Dis. 2021;17(1):29–32.

7. Lambert JE, Ramos-Roman MA, Browning JD, Parks EJ. Increased de novo lipogenesis is a distinct characteristic of individuals with nonalcoholic fatty liver disease.. Gastroenterology 2014;146(3):726–35.

8. Fletcher JA et al. Impaired ketogenesis and increased acetyl-CoA oxidation promote hyperglycemia in human fatty liver. JCI Insight 2019;4(11). doi:10.1172/jci.insight.127737

9. White PJ et al. The BCKDH Kinase and Phosphatase Integrate BCAA and Lipid Metabolism via Regulation of ATP-Citrate Lyase.. Cell Metab. 2018;27(6):1281–1293.e7.

10. White PJ, Newgard CB. Branched-chain amino acids in disease. Science (80-.). 2019;363(6427):582–583.

11. White PJ et al. The BCKDH Kinase and Phosphatase Integrate BCAA and Lipid Metabolism via Regulation of ATP-Citrate Lyase. Cell Metab. [published online ahead of print: May 11, 2018]; doi:10.1016/j.cmet.2018.04.015

12. Potapova IA, El-Maghrabi MR, Doronin S V., Benjamin WB. Phosphorylation of Recombinant Human ATP:Citrate Lyase by cAMP-Dependent Protein Kinase Abolishes Homotropic Allosteric Regulation of the Enzyme by Citrate and Increases the Enzyme Activity. Allosteric Activation of ATP:Citrate Lyase by Phosphorylated Sug. Biochemistry 2000;39(5):1169–1179.

13. Martinez Calejman C et al. mTORC2-AKT signaling to ATP-citrate lyase drives brown adipogenesis and de novo lipogenesis. Nat. Commun. 2020;11(1):1–16.

14. Romeo S et al. Genetic variation in PNPLA3 confers susceptibility to nonalcoholic fatty liver disease. Nat. Genet. 2008;40(12):1461–1465.

15. Tang JJ et al. Inhibition of SREBP by a small molecule, betulin, improves hyperlipidemia and insulin resistance and reduces atherosclerotic plaques. Cell Metab. 2011;13(1):44–56.

16. Hawkins JL et al. Pharmacologic inhibition of site 1 protease activity inhibits sterol regulatory element-binding protein processing and reduces lipogenic enzyme gene expression and lipid synthesis in cultured cells and experimental animals. J. Pharmacol. Exp. Ther. 2008;326(3):801–808.

17. Newgard CB et al. A branched-chain amino acid-related metabolic signature that differentiates obese and lean humans and contributes to insulin resistance.. Cell Metab. 2009;9(4):311–26.

18. Wang TJ et al. Metabolite profiles and the risk of developing diabetes.. Nat. Med. 2011;17(4):448–53.

19. Palmer ND et al. Metabolomic Profile Associated With Insulin Resistance and Conversion to Diabetes in the Insulin Resistance Atherosclerosis Study. J. Clin. Endocrinol. Metab. 2015;100(3):E463–E468.

20. Tai ES et al. Insulin resistance is associated with a metabolic profile of altered protein metabolism in Chinese and Asian-Indian men.. Diabetologia 2010;53(4):757–67.

21. Lotta LA et al. Genetic Predisposition to an Impaired Metabolism of the Branched-Chain Amino Acids and Risk of Type 2 Diabetes: A Mendelian Randomisation Analysis.. PLoS Med. 2016;13(11):e1002179.

22. Goffredo M et al. A Branched-Chain Amino Acid-Related Metabolic Signature Characterizes Obese Adolescents with Non-Alcoholic Fatty Liver Disease. Nutrients 2017;9(7). doi:10.3390/NU9070642

23. Gaggini M et al. Altered amino acid concentrations in NAFLD: Impact of obesity and insulin resistance. Hepatology 2018;67(1):145–158.

24. van den Berg EH et al. Non-Alcoholic Fatty Liver Disease and Risk of Incident Type 2 Diabetes: Role of Circulating Branched-Chain Amino Acids. Nutrients 2019;11(3). doi:10.3390/NU11030705

25. Grzych G et al. Plasma BCAA Changes in Patients With NAFLD Are Sex Dependent. J. Clin. Endocrinol. Metab. 2020;105(7):2311–2321.

26. Lischka J et al. A branched-chain amino acid-based metabolic score can predict liver fat in children and adolescents with severe obesity. Pediatr. Obes. 2021;16(4). doi:10.1111/IJPO.12739

27. Felig P, Marliss E, Cahill GF. Plasma amino acid levels and insulin secretion in obesity.. N. Engl. J. Med. 1969;281(15):811–6.

28. Mahendran Y et al. Genetic evidence of a causal effect of insulin resistance on branched-chain amino acid levels. Diabetologia 2017;60(5):873–878.

29. Ridaura VK et al. Gut microbiota from twins discordant for obesity modulate metabolism in mice.. Science 2013;341(6150):1241214.

30. Pedersen HK et al. Human gut microbes impact host serum metabolome and insulin sensitivity. Nature 2016;535(7612):376–381.

31. Herman MA, She P, Peroni OD, Lynch CJ, Kahn BB. Adipose tissue branched chain amino acid (BCAA) metabolism modulates circulating BCAA levels.. J. Biol. Chem. 2010;285(15):11348–56.

32. She P et al. Obesity-related elevations in plasma leucine are associated with alterations in enzymes involved in branched-chain amino acid metabolism.. Am. J. Physiol. Endocrinol. Metab. 2007;293(6):E1552–63.

33. Shin AC et al. Brain Insulin Lowers Circulating BCAA Levels by Inducing Hepatic BCAA Catabolism.. Cell Metab. [published online ahead of print: October 7, 2014]; doi:10.1016/j.cmet.2014.09.003

34. Lian K et al. Impaired adiponectin signaling contributes to disturbed catabolism of branched-chain amino acids in diabetic mice.. Diabetes 2015;64(1):49–59.

35. Sweatt AJ et al. Branched-chain amino acid catabolism: unique segregation of pathway enzymes in organ systems and peripheral nerves. Am. J. Physiol. Metab. 2004;286(1):E64–E76.

36. Neinast MD et al. Quantitative Analysis of the Whole-Body Metabolic Fate of Branched-Chain Amino Acids. Cell Metab. 2019;29(2):417–429.e4.

37. White PJ et al. Branched-chain amino acid restriction in Zucker-fatty rats improves muscle insulin sensitivity by enhancing efficiency of fatty acid oxidation and acyl-glycine export. Mol. Metab. 2016;5(7):538–551.

38. Patel MJ et al. Race and Sex Differences in Small-Molecule Metabolites and Metabolic Hormones in Overweight and Obese Adults. Omi. A J. Integr. Biol. 2013;17(12):627–635.

39. Newbern D et al. Sex Differences in Biomarkers Associated With Insulin Resistance in Obese Adolescents: Metabolomic Profiling and Principal Components Analysis. J. Clin. Endocrinol. Metab. 2014;99(12):4730–4739.

40. Perng W, Rifas-Shiman SL, Hivert M-F, Chavarro JE, Oken E. Branched Chain Amino Acids, Androgen Hormones, and Metabolic Risk Across Early Adolescence: A Prospective Study in Project Viva.. Obesity (Silver Spring). 2018;26(5):916–926.

41. Saito Y et al. LLGL2 rescues nutrient stress by promoting leucine uptake in ER+ breast cancer. Nature 2019;569(7755):275–279.

42. Mardinoglu A et al. Genome-scale metabolic modelling of hepatocytes reveals serine deficiency in patients with non-alcoholic fatty liver disease. Nat. Commun. 2014;5(1):3083.

43. Mardinoglu A et al. Personal model assisted identification of NAD <sup>+</sup> and glutathione metabolism as intervention target in NAFLD. Mol. Syst. Biol. 2017;13(3):916.

44. White PJ et al. Muscle-Liver Trafficking of BCAA-Derived Nitrogen Underlies Obesity-Related Glycine Depletion. Cell Rep. 2020;33(6) 108375.

45. Innes JK, Calder PC. Omega-6 fatty acids and inflammation. Prostaglandins, Leukot. Essent. Fat. Acids 2018;132:41–48.

46. Brunt EM, Janney CG, Di Bisceglie AM, Neuschwander-Tetri BA, Bacon BR. Nonalcoholic steatohepatitis: A proposal for grading and staging the histological lesions. Am. J. Gastroenterol. 1999;94(9):2467–2474.

47. Chella Krishnan K et al. Liver Pyruvate Kinase Promotes NAFLD/NASH in Both Mice and Humans in a Sex-Specific Manner. Cell. Mol. Gastroenterol. Hepatol. 2021;11(2):389–406.

48. Hui ST et al. The Genetic Architecture of Diet-Induced Hepatic Fibrosis in Mice. Hepatology 2018;68(6):2182–2196.

49. Xiong X et al. Landscape of Intercellular Crosstalk in Healthy and NASH Liver Revealed by Single-Cell Secretome Gene Analysis. Mol. Cell 2019;75(3):644–660.e5.

50. Stöckli J et al. Metabolomic analysis of insulin resistance across different mouse strains and diets. J. Biol. Chem. 2017;292(47):19135–19145.

51. León-Mimila P et al. Genome-Wide Association Study Identifies a Functional SIDT2 Variant Associated With HDL-C (High-Density Lipoprotein Cholesterol) Levels and Premature Coronary Artery Disease. Arterioscler. Thromb. Vasc. Biol. 2021;41(9):2494–2508.

52. Kim D et al. TopHat2: accurate alignment of transcriptomes in the presence of insertions, deletions and gene fusions. Genome Biol. 2013;14(4). doi:10.1186/GB-2013-14-4-R36

53. Trapnell C et al. Transcript assembly and quantification by RNA-Seq reveals unannotated transcripts and isoform switching during cell differentiation.. Nat. Biotechnol. 2010;28(5):511–5.

54. Gerstein MB et al. Architecture of the human regulatory network derived from ENCODE data. Nature 2012;489(7414):91–100.

