## Supplemental Materials for "Altered branched-chain α-keto acid metabolism is a feature of NAFLD in individuals with severe obesity"

**Table S1. PCA Factors**

| PCA Factor | Metabolite Components |
|---|---|
| **Factor 1 – Even chain acylcarnitines** | C14:1, C12:1, C2, C16:1,C14:2,C4-OH, C16:2,C14:1-OH, C12, C18:1, C6-DC/C8-OH, C16:1-OH/C14:1-DC, C14, C12-OH/C10-DC, C18:1-OH/C16:1-DC, C16, C10:1, C10, C14-OH/C12-DC, C18:2, C8, C8:1-DC, C18:1-DC |
| **Factor 2 – Glucosylceramides** | Glucosylceramides d18:1/C24, d18:1/C16, d18:1/C24:1, d18:1/C22, d18:1/C23, d18:1/C18, d18:1/C20; ceramide C16 |
| **Factor 3 – Amino acid related** | Phenylalanine, leucine/isoleucine, methionine, arginine, tyrosine, valine, ornithine, C3 acylcarinitine, proline, citrulline, KMV, histidine |
| **Factor 4 – Ceramides** | Ceramides C26:1, C20, C26, C25, C18; glucosylceramide d18:1/C26 |
| **Factor 5 – Branched-chain keto and amino acids** | KIC, KMV, KIV, leucine/isoleucine, valine |
| **Factor 6 – Medium chain OH/DC acylcarnitines** | C8:1-OH/C6:1-DC, C10-OH/C8-DC, C8:1-DC |
| **Factor 7 – Glycine related amino acids** | Glycine, serine, histidine |
| **Factor 8 – C18/C16 acylcarnitines** | C18, C16, C18:2, C18:1 |
| **Factor 9 – Long chain ceramides** | Ceramides C24:1, C16, C20:1, C18 |
| **Factor 10 – Long chain acylcarnitines** | C20:4, C22, C18:2-OH |
| **Factor 11 – Medium chain unsaturated acylcarnitines** | C8:1, C10:3, C10:2 |
| **Factor 12 – Long chain ceramides** | Ceramides C23, C22, C24 |
| **Factor 13 – Hydroxyisovaleryl/malonyl carnitine** | C5-OH/C3-DC |
| **Factor 14 – Alanine, proline** | Alanine, proline |
| **Factor 15 – C20 acylcarnitine** | C20 |
| **Factor 16 – Short chain acylcarnitines** | C4/Ci4, C3, C5 |
| **Factor 17 – Long chain dicarboxyl acylcarnitines** | C20-OH/C18-DC, C18:1-DC |
| **Factor 18 – Medium chain acylcarnitines** | C8, C10, C10:1 |

PCA factors were created using 80 metabolites. Top 18 factors displayed above (each with eigenvalue >1) explain 73% of total variance.

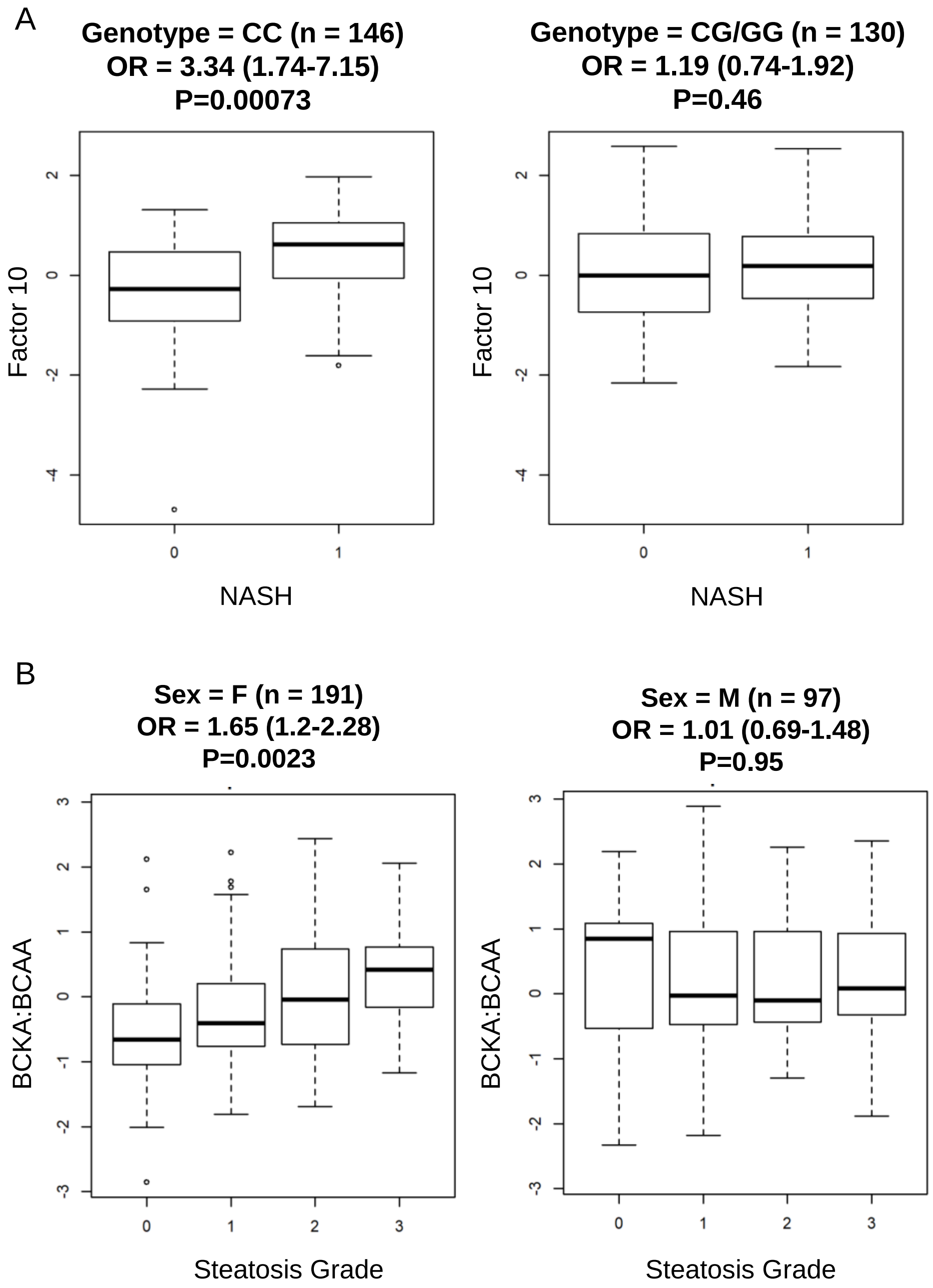


**Figure S1. Genotype and sex interactions.** Panel A shows association of Factor 10 with NASH status is not present in carriers of the Ile148Met G allele of *PNPLA3*. Panel B shows association of BCKA:BCAA ratio with steatosis grade is present in females but not males.

**Table S2. Baseline characteristics of the gene expression cohorts**

| Trait | QHLI (N = 60) | MOBES (N = 107) |
|---|---|---|
| Age (Y) | 45.0 (40.0 - 51.0) | 35.0 (28.0 - 44.5) |
| Female (%) | 42 (70) | 93 (86.9) |
| BMI (kg/m2) | 49.7 (46.6 - 52.7) | 43.2 (39.1 - 48.5) |
| Glucose (mg/dL) | 117.0 (99.0 - 146.7) | 91.0 (80.3 - 99.8) |
| TG (mg/dL) | 129.3 (100.7 - 159.0) | 124.0 (91.0 - 162.0) |
| TC (mg/dL) | 181.3 (162.3 - 203.5) | 172.0 (147.0 - 194.0) |
| HDL-C (mg/dL) | 48.7 (42.4 - 58.8) | 36.0 (31.0 - 43.0) |
| LDL-C (mg/dL) | 106.0 (83.2 - 121.0) | 105.0 (86.6 - 129.0) |
| ALT (UI/L) | 29.5 (21.0 - 41.0) | 27.0 (21.0 - 38.0) |
| AST (UI/L) | 23.0 (18.0 - 31.0) | 26.0 (21.0 - 32.0) |
| GGT (UI/L) | 29.0 (22.5 - 47.0) | 19.0 (14.0 - 26.0) |

Data are shown as median (interquartile range) or n (%). Abbreviations: BMI, Body Mass Index; TG, Triglycerides; TC, Total Cholesterol; HDL-C, high-density lipoprotein cholesterol; LDL-C, low-density lipoprotein; ALT, alanine aminotransferase; AST, aspartate aminotransferase; GGT, gamma-glutamyl transferase.

**Table S3. Primers used for qPCR**

| Target | Forward Primers | Reverse Primers |
|---|---|---|
| **h*BCKDK*** | 5’ -TGAGAAGTGGGTGGACTTTGC- 3’ | 5’ -ATGGCATTCTTGAGCAGCTC- 3’ |
| **m*Bckdk*** | 5’ -ATCTGTACTCGTCTGTCGCC- 3’ | 5’ -TCCGTTGATGCGGACTCG- 3’ |
| **m*Fasn*** | 5’ -AGTCAGCTATGAAGCAATTGTGGA- 3’ | 5’ -CACCCAGACGCCAGTGTTC- 3’ |
| ***RPLP0*** | 5’ -TCTGCATTCTCGCTTCCTGG- 3’ | 5’-CCAGGACTCGTTTGTACCCG- 3’ |
